# Evaluating an Adjusting Alcohol Purchase Task as a Brief Measure of Behavioural Economic Demand for Alcohol in a Large Sample of Community Adults

**DOI:** 10.64898/2026.07.02.26357139

**Authors:** Sophie G. Coelho, Kyla L. Belisario, Matthew T. Keough, James MacKillop

**Author notes:** Please address correspondence to: James MacKillop, Peter Boris Centre for Addictions Research, McMaster University/St. Joseph’s Healthcare Hamilton, 100 West 5th Street, Hamilton, Ontario L8P 3R2, Canada.

## Abstract

Alcohol demand is commonly assessed using hypothetical alcohol purchase tasks (APTs), from which individual demand curves are constructed and yield multiple indices of reinforcing value. Procedurally, APTs can confer participant burden, and existing brief alternatives cannot produce demand curves or derived indices. Thus, we evaluated a novel, adjusting APT that efficiently and idiographically assesses alcohol demand while preserving the benefits of a full task. Adults reporting past-six-month alcohol use (*n*=897) completed either the adjusting or full APT, the former utilizing a binary-search-style algorithm to administer six prices from the full APT’s price set based on level of alcohol demand. The adjusting APT reduced item burden by 49% and produced well-fitting individual demand curves. Average demand intensity and elasticity estimates did not differ significantly by modality, whereas *O*_max_ and breakpoint estimates were significantly higher on the adjusting APT, though only by $3 each. All demand indices from both APTs were positively associated with alcohol use and problems, with similar magnitude by modality. Results provide support for the adjusting APT as a brief measure of alcohol demand that retains demand-curve-based indices of reinforcing value.

## Introduction

Behavioural economic accounts of alcohol use emphasize the relative reinforcing value of alcohol as a central determinant of drinking behaviour (Acuff et al., 2023; Bickel et al., 2014; Hursh et al., 2005; MacKillop, 2016). The relative reinforcing value of alcohol can be operationalized as behavioural economic demand for alcohol, describing alcohol consumption as a function of its cost (Hursh, 1980; Hursh et al., 2005; Hursh & Silberberg, 2008). Demand curve analyses yield multiple indices quantifying the consumption-cost relation, including consumption at zero cost (demand intensity), maximum expenditure across the demand curve (*O*_max_), the price that suppresses consumption to zero (breakpoint), and the extent to which consumption decreases as a function of escalating price (price sensitivity). These indices are related, yet topographically distinct, corresponding to different parts of the demand curve and requiring varying levels of cost-benefit decision-making (Amlung et al., 2024; MacKillop, 2016; MacKillop et al., 2014). Alcohol demand is most commonly assessed using the Alcohol Purchase Task (APT; Kaplan et al., 2019; Murphy & MacKillop, 2006), which is a hypothetical purchase task wherein individuals estimate how much alcohol they would consume across escalating prices. Meta-analyses have found robust associations of individual differences in demand indices from the APT with individual differences in the quantity and frequency of alcohol use, heavy drinking, alcohol-related problems, and hazardous alcohol use (Martínez-Loredo et al., 2021). Meta-analytic research also supports the sensitivity of demand indices from the APT to state manipulations, including alcohol cue exposure, negative affect induction, pharmacological interventions, and opportunity cost (Acuff et al., 2020). Further, the APT has been applied in clinical and policy research, for example in forecasting brief intervention responses (e.g., Cassidy et al., 2019; MacKillop & Murphy, 2007; Murphy et al., 2015) and in simulating impacts of low alcohol prices on heavy drinking (Murphy & MacKillop, 2006), respectively. This prior research collectively supports the APT’s utility as a versatile measure of trait and state incentive salience of alcohol.

The APT typically requires participants to report hypothetical alcohol consumption across 20 to 30 prices, posing potential burden to participants when administered within large batteries of measures, intensive longitudinal studies with frequent assessment, or paradigms requiring administration of multiple versions of the task (e.g., examining the behavioural economic substitutability of different commodities). Thus, streamlining the APT is essential to broadening its use. An initial abbreviation of the APT resulted in the Brief Assessment of Alcohol Demand (BAAD; Owens et al., 2015), which consists of just three face-valid items, each explicitly assessing an index of alcohol demand (intensity, *O*_max_, or breakpoint). Prior research has supported the BAAD’s concurrent and convergent validity (Amlung et al., 2015; Aston & Merrill, 2023; Hardy et al., 2021; Merrill & Aston, 2020; Owens et al., 2015), but the BAAD only captures demand indices that can be assessed explicitly (as opposed to implicitly on the traditional measure) and individual demand curves cannot be estimated, thus meaning elasticity cannot be derived. This limits the BAAD’s use given that elasticity is robustly associated with alcohol use outcomes both cross-sectionally and longitudinally (Bird et al., 2024; Martínez-Loredo et al., 2021). Further, elasticity, as well as *O*_max_ and breakpoint when assessed using the APT, is relatively less subject to conscious reflective cognitive processing and putatively less susceptible to bias (Acker & MacKillop, 2013).

A brief, adjusting APT was recently developed to address limitations of the BAAD, assessing alcohol demand in just six items while still permitting demand curve estimation (Coelho et al., 2026). The adjusting APT is an idiographic measure, employing a binary-search-style algorithm to administer tailored sequences of six price points contingent on the individual’s level of alcohol demand. This adjusting algorithm, in utilizing the full price range of the APT, circumvents the trade-off between adequate measurement of lower versus higher levels of alcohol demand applicable to abbreviated fixed-item APTs. Namely, eliminating low prices risks increased non-random missing data should participants with the lowest demand no longer report non-zero consumption at the two or more prices needed to calculate elasticity. Alternatively, eliminating high prices risks ceiling effects, particularly for breakpoint and *O*_max_ should participants with the highest demand no longer reach the prices that respectively would have suppressed consumption to zero and maximized expenditure. Overcoming this trade-off is important as an optimal fixed subset of prices may be sample-specific, contingent on characteristics such as income, regional alcohol prices, or level of alcohol use.

The adjusting APT was initially evaluated in two independent samples of heavy-drinking young adults by using the adjusting APT algorithm to retrospectively infer participants’ adjusting APT responses from their responses on a full-length, fixed-item APT (Coelho et al., 2026). In both samples, individual demand curves from the adjusting APT fit the data well, and demand indices estimated from the adjusting APT corresponded to those estimated from the full APT. Further, demand indices from the adjusting APT were consistently associated with alcohol use and problems, with similar effect sizes to corresponding associations using demand indices from the full APT, supporting convergent validity. These findings provide initial support for the adjusting APT as a multidimensional measure of alcohol demand, though there remains a need for further examination of its psychometric properties following stand-alone administration. As previous studies have only inferred participants’ adjusting APT responses retrospectively, participants still reported their consumption across prices presented in ascending order, rather than in descending order or non-monotonically as several adjusting APT permutations require. Thus, there is a need to examine whether demand indices estimated from the adjusting and full APTs remain comparable with the former administered according to its algorithm. Further, the adjusting APT has been evaluated only among heavy-drinking young adults. Extending the use of this measure to community samples reporting alcohol use is important to establish its validity across a wider spectrum of ages and drinking levels.

The current study aimed to evaluate a brief six-item adjusting APT in a large sample of community adults reporting alcohol use. Participants were randomly assigned to complete either an adjusting or full APT. We examined whether observed (intensity, *O*_max_, breakpoint) and derived (elasticity) indices of alcohol demand could be validly estimated from the adjusting APT data, and the extent to which these demand indices were comparable to those estimated from an independently administered full APT. We also assessed the convergent validity of demand indices obtained from the adjusting APT by estimating their associations with alcohol use and problems, and we examined whether these associations were of comparable magnitude to associations of demand indices obtained from the full APT with these outcomes. We hypothesized that the adjusting and full APTs would produce similar mean levels of demand across indices, and that demand indices from both APTs would be associated with alcohol use and problems.

## Method

### Participants and Procedures

Data were from a longitudinal observational cohort study of health behaviour in community adults. The cohort was recruited from an existing research registry of community-dwelling adults in [MASKED FOR REVIEW]. Cohort eligibility criteria included being between 18 and 65 years of age at baseline, minimum ninth-grade education, being willing to consider participating in research, and having no diagnosed terminal illness. Eligible participants completed an extended in-person assessment upon enrollment in mid-September to mid-October of 2018 and subsequently completed sixteen follow-up assessments over the following seven years. Participants completed assessments online via the web-based Research Electronic Data Capture (REDCap) software (Harris et al., 2019). Assessments each included five quality control items with unambiguously correct responses to confirm adequate attention and effort, and data were retained only when three or more of these items were answered correctly.

Analyses reported herein used data from an assessment wave in October, 2025, which was when the adjusting APT evaluation protocol was implemented. Of the initial 1502 participants enrolled in the cohort, 1187 participants (79.03%) provided data that met quality control criteria at this wave. The 897 of these participants (75.57%) reporting past-six-month alcohol use were randomly assigned to complete either the adjusting APT (*n* = 435) or full APT (*n* = 462) and formed this study’s sample. At baseline, participants in the current sample were significantly older, more likely to have obtained at least a bachelor’s degree, and more likely to report higher subjective financial status compared to participants from the larger cohort who were excluded from the current sample (*n* = 605); no other significant baseline demographic differences between these groups were observed (see Table S1).

## Measures

### Demographic Characteristics

At baseline, participants reported their age, sex, gender, race, subjective financial status, annual household income, highest level of education, and employment status. Response options for demographic items are shown in Table 1.

**Table 1.** Sample characteristics (*n* = 897)

|  | *n* | % |
| --- | --- | --- |
| Sex: female | 553 | 61.65 |
| Gender |  |  |
| Man | 340 | 37.90 |
| Woman | 541 | 60.31 |
| Transgender/nonbinary | 16 | 1.78 |
| Race |  |  |
| Multiple | 36 | 4.01 |
| White | 715 | 79.71 |
| Black | 16 | 1.78 |
| East Asian | 38 | 4.24 |
| South Asian | 33 | 3.68 |
| Southeast Asian | 6 | 0.67 |
| Middle Eastern | 13 | 1.45 |
| First Nations/Indigenous | 6 | 0.67 |
| Pacific Islander | 3 | 0.33 |
| Other | 31 | 3.46 |
| Latinx: yes | 35 | 3.90 |
| Subjective financial status |  |  |
| Not enough to pay some bills no matter how hard you try | 35 | 3.90 |
| Enough to pay bills, but have had to cut back | 193 | 21.52 |
| Enough to pay bills without cutting back but no “extras” | 232 | 25.86 |
| Enough money for “extras” | 437 | 48.72 |
| Annual household income |  |  |
| < \$30,000 | 54 | 6.02 |
| \$30,000 to < \$60,000 | 112 | 12.49 |
| \$60,000 to < \$90,000 | 162 | 18.06 |
| \$90,000 to < \$120,000 | 187 | 20.85 |
| \$120,000 to < \$150,000 | 110 | 12.26 |
| ≥ \$150,000 | 272 | 30.32 |
| Highest level of education |  |  |
| High school graduate (or GED) or less | 47 | 5.24 |
| Some college/university | 212 | 23.63 |
| Associates degree | 85 | 9.48 |
| Bachelor’s degree | 309 | 34.45 |
| Master’s or professional degree | 244 | 27.20 |
| Employment Status: employed (part-time or full-time) | 687 | 76.59 |
|  | *M* | *SD* |
| Baseline age | 42.86 | 14.46 |
| Drinks/week | 4.71 | 6.03 |
| Drinking days/week | 2.21 | 1.80 |
| HDD/week | 0.29 | 1.01 |
| AUDIT total score | 3.74 | 3.79 |

*Note.* *M* = mean; *SD* = standard deviation; *df* = degrees of freedom; HDD = heavy drinking days; AUDIT = Alcohol Use Disorder Identification Test.

### Alcohol Demand

#### Full Alcohol Purchase Task

The full APT (Murphy & MacKillop, 2006) asked participants to report the number of standard drinks that they would consume in a typical drinking scenario (i.e., what, where, and with whom they typically drink) at each of 19 drink prices. Drink prices ranged from $0 to $40 to reflect typical drink prices where the study was conducted, and responses were capped at 99 drinks for biological plausibility. Task instructions outlined several assumptions, including that drinks purchased were for personal consumption, and that drinking before or after the drinking scenario and stockpiling drinks for future consumption were prohibited. Complete task instructions and price structure are in the Supplemental Materials. The APT was administered in blocks of five (four for the last four prices), such that when participants reported that they would consume zero standard drinks at any price within a given block, subsequent blocks were not administered to minimize participant burden.

#### Adjusting Alcohol Purchase Task

The adjusting APT (Coelho et al., 2026) was consistent with the full APT in its instructions, but used a branching item selection algorithm to provide each participant with a tailored subset of five to six prices sampled from a range of prices. There were 30 prices from which personalized price sequences were selected, ranging from $0 to $40 like the full APT but with greater granularity between $0 and $15. The adjusting APT’s binary-search-style algorithm involved repeatedly halving the search space (i.e., all non-zero prices) with participants’ directional cues, defined as their reports of zero (lower) versus non-zero (higher) consumption, determining the personalized price sequence presented. This algorithm yielded 16 permutations of price sequences, each four to five non-zero prices in length. The adjusting APT algorithm is presented in Figure 1, and complete task instructions and price structure are in the Supplemental Materials. Practically, participants first reported the number of drinks that they would consume if alcohol were free, as this value is required to estimate intensity (*Q*_0_) and elasticity (α). They next reported the number of drinks that they would consume at $11 per drink, which is the median price of the full set of non-zero prices. Reporting zero consumption at this price would prompt subsequent administration of a lower price ($3, the median between the first non-zero price and $11) whereas reporting non-zero consumption at this price would prompt subsequent administration of a higher price ($22, the median between $11 and the highest price). Subsequent items were administered following this pattern of administering a lower price following zero consumption or a higher price following non-zero consumption, for each participant yielding reports of their hypothetical alcohol use across a personalized sequence of five to six prices (including $0). The frequencies of administration of each adjusting APT permutation are reported in Table S3.

**Figure 1.**
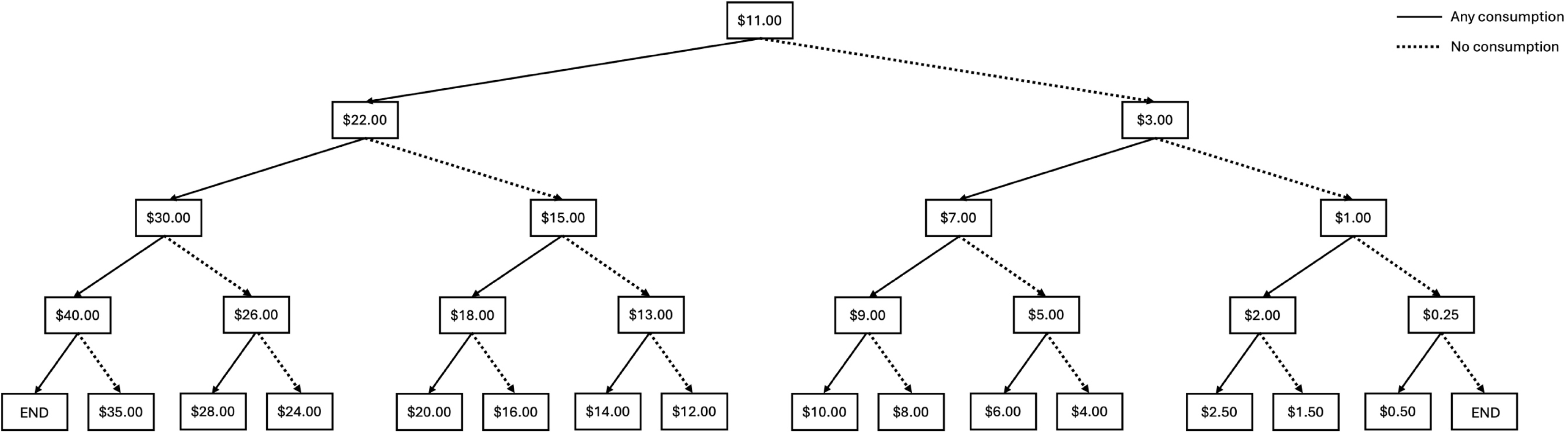
Adjusting alcohol purchase task algorithm. Adjusting alcohol purchase task algorithm. All participants first reported the number of standard drinks they would consume if alcohol were free.

**Figure 2.**
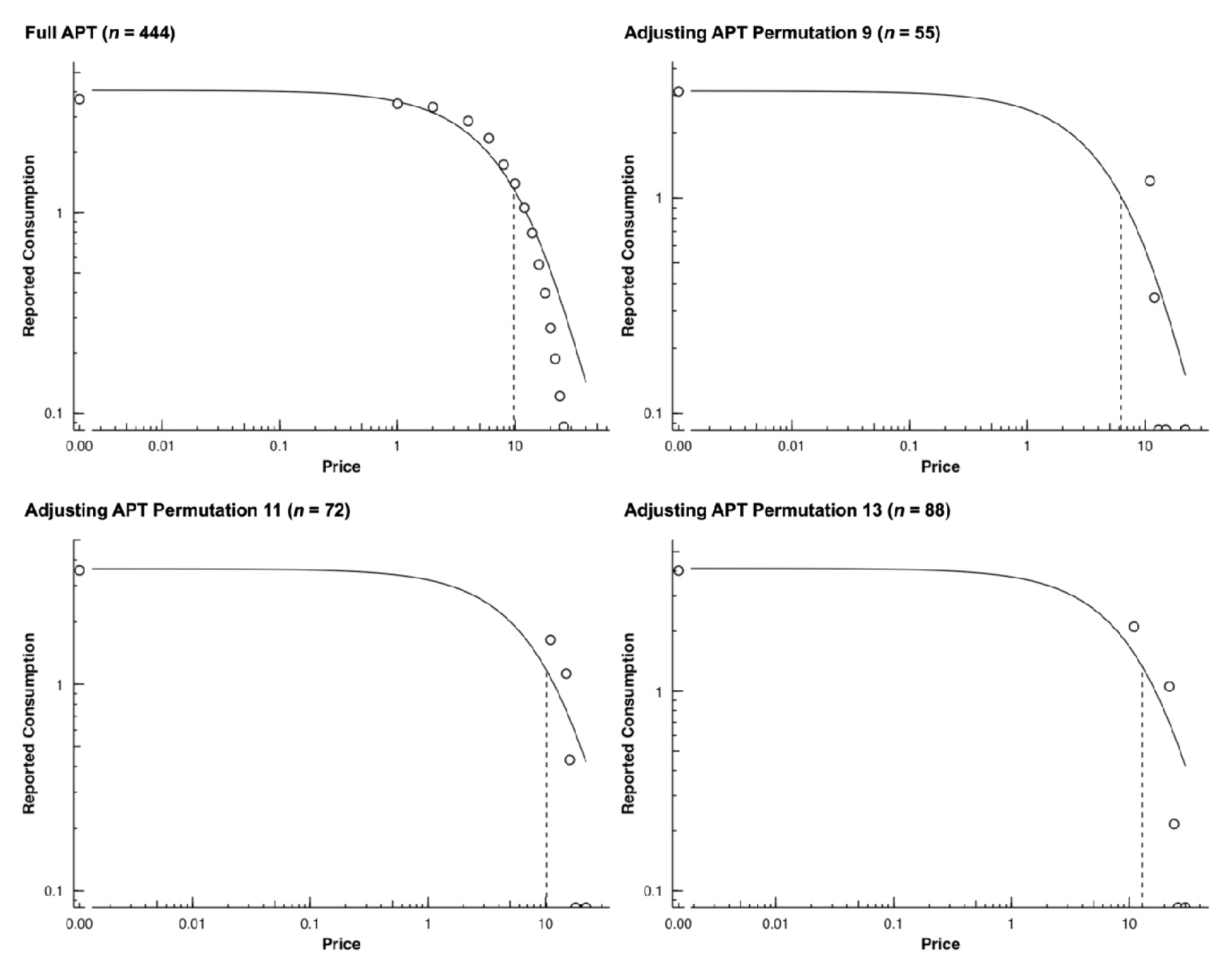
Mean Aggregate Demand Curves for the Full Alcohol Purchase Task and the Three Most Frequent Permutations of the Adjusting Alcohol Purchase Task. Mean aggregate demand curves for the full alcohol purchase task and the three most frequent permutations of the adjusting alcohol purchase task. APT = alcohol purchase task; *n* = number of participants for whom individual demand curves could be estimated (i.e., provided systematic data and reported alcohol consumption at two or more prices) and are thus represented in the plot for a given version of the alcohol purchase task. Both price (*x* axis) and reported consumption (*y* axis) are presented in log scale. Median *R*^2^ values denoting fit of the individual demand curves were 0.87 for the full APT, 0.81 for permutation 9 of the adjusting APT, 0.80 for permutation 11 of the adjusting APT, and 0.84 for permutation 13 of the adjusting APT. Plots for all permutations of the adjusting APT are provided in Figure S1, and price sequences corresponding to each adjusting APT permutation are reported in Table S3.

### Alcohol Use and Problems

Participants completed the Daily Drinking Questionnaire (DDQ; Collins et al., 1985), on which they reported the total number of standard drinks (1.5 fl. oz. of distilled spirits, 5 fl. oz. of wine, or 12 fl. oz. of beer) consumed each day of the week during a typical week in the past month. We calculated the total number of standard drinks consumed, drinking days, and heavy drinking days (HDD; at least four [female participants] or five [male participants] standard drinks) per typical week in the past month. For analyses, number of HDD per typical week was dichotomized as any versus no HDD, as only a minority of participants (*n* = 109, 12.15%) reported any HDD per typical week, among whom a majority reported just one (*n* = 41, 37.61%) or two (*n* = 34, 31.19%) HDD. Participants also completed the Alcohol Use Disorder Identification Test (Saunders et al., 1993), consisting of 10 items assessing alcohol-related behaviours and problems. Items are rated on a five-point (first eight items) or three-point (last two items) ordinal scale from 0 to 4 and are summed to obtain a continuous index of hazardous use. The AUDIT exhibited acceptable internal consistency in this sample (*ω* = 0.86).

## Data Analysis

### Processing Demand Data and Estimating Demand Indices

The APT data were processed and demand indices were estimated using the *beezdemand* package in R (Kaplan et al., 2019; R Core Team, 2022). Observations exhibiting one or more signs of non-systematic responding (Kaplan et al., 2019; Stein et al., 2015) were removed, with signs including: (i) reversals from zero, defined as any non-zero consumption following any instance of zero consumption (the most conservative application of this criterion); (ii) price-to-price increases in consumption at more than 10% of price increments (i.e., “bounces”); and (iii) constant or increasing consumption across escalating prices, indicated by a less than 0.025 log-unit reduction in consumption per log-unit in price. Demand indices estimated for use in analyses were consumption at zero cost (intensity), maximum expenditure on alcohol (*O*_max_), the price at which alcohol consumption was suppressed to zero (breakpoint), and sensitivity of consumption to price (i.e., the aggregate slope of the demand curve, elasticity). Intensity, *O*_max_, and breakpoint were estimated empirically, obtained directly from participants’ responses. When consumption was never suppressed to zero, breakpoint was coded as $45 to represent the preceding price increment ($5) higher than the highest price ($40). Elasticity was derived from the individual demand curves, which were estimated using the exponentiated demand curve equation (Koffarnus et al., 2015): 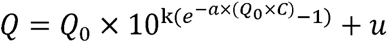, where *Q* = consumption at each price; *Q*_0_ = derived intensity, or consumption at zero cost; *k* = the span of the demand curve, constant across individuals; α = the rate of decline in consumption with increasing price (i.e., elasticity); *C* = the cost of the commodity; and *u =* residuals. Elasticity was not estimated for participants reporting no consumption across all prices or consumption only when alcohol was free, as its estimation requires two points of demand (i.e., non-zero consumption at two or more prices). Demand curves were estimated and plotted separately for each permutation of the adjusting APT and the full APT given differences in the prices used to estimate demand curves, though with a common *k* value to permit analyses of individual differences in elasticity. This *k* was the log_10_-transformed mean consumption at the highest price ($40) subtracted from the log_10_-transformed mean consumption at the lowest price ($0) added to 0.5 (Kaplan et al., 2019), calculated separately for the full APT and adjusting APT and then averaged to obtain a common *k* value.

### Analyses

Prior to analyses, extreme outliers in demand indices, defined as values more than 3.99 standard deviations higher or lower than the mean, were winsorized to one unit (intensity, *O*_max_, breakpoint) or 0.001 units (elasticity) higher or lower than the next non-outlying value, separately for each version of the APT. Extreme outliers in number of standard drinks per typical week were also winsorized to one unit higher than the next non-outlying value.

To evaluate the extent to which the adjusting APT obtained systematic responses (i.e., decreasing consumption with increasing price) despite its descending and non-monotonic price sequences, we examined rates of non-systematic response signs among participants who completed each version of the APT. We also examined the proportion of participants for whom elasticity could not be estimated (i.e., due to consumption reported at fewer than two prices) and who did not reach breakpoint (i.e., due to reporting non-zero consumption at the highest price). To evaluate the suitability of adjusting APT data for demand curve analyses, we examined fit of the demand curves from both the adjusting and full APTs using *R*^2^ values for the individual demand curves and plots of the mean aggregate demand curves (separate plots for each adjusting APT permutation); well-fitting demand curves supported the validity of derived elasticity estimates. We compared the average number of items administered and average task duration between the full and adjusting APTs to empirically evaluate the relative efficiency of the adjusting APT. Further, we examined the proportion of those who completed the adjusting APT whose breakpoint values corresponded to prices not available on the full APT to assess the incremental value of the increased price granularity afforded by the adjusting APT’s algorithm.

Next, we obtained descriptive statistics on demand indices estimated from both the adjusting and full APTs, and unadjusted estimates of differences in these demand indices between participants who completed the different APT versions. As a sensitivity analysis, group differences in demand indices were additionally examined by regressing each demand index on a dichotomous indicator of APT version, adjusting for several demographic and alcohol-related covariates (age, sex, race, subjective financial status, highest level of education, employment status, hazardous alcohol use); however, adjusted group differences did not differ substantively from unadjusted group differences and thus the latter were retained for parsimony. As participants were randomly assigned to complete either the adjusting or full APT, group differences in demand indices should ostensibly reflect differences between the two tasks.

Finally, we assessed the convergent validity of the adjusting APT, and the extent to which its convergent validity differed from that of the full APT. To this end, we estimated regression models with each of the following dependent variables regressed on demand indices: number of standard drinks per typical week, number of drinking days per typical week, likelihood of any HDD per typical week, and level of hazardous alcohol use (i.e., AUDIT total scores). Number of standard drinks and number of drinking days were modelled as count outcomes using a negative binomial distribution (to accommodate overdispersion) and log link function, likelihood of any HDD was modelled as a binary outcome using a binomial distribution and logit link function, and hazardous alcohol use was modelled as a continuous outcome using a gaussian distribution and identity link function. Separate models were estimated for each demand index and demand indices were standardized prior to estimating models for interpretability. Each model also included an interaction between the demand index and the APT version from which it was obtained. We were interested both in whether demand indices from the adjusting APT were associated with alcohol use and problems, and whether associations of demand indices with alcohol use and problems differed between the adjusting and full APTs. To this end, we obtained associations of demand indices with alcohol use and problems conditional on each of the adjusting and full APTs by changing the reference category for the APT version indicator variable. Differences in associations of demand indices with alcohol use and problems between the adjusting and full APTs were indicated by statistically significant interactions between demand indices and APT version. All models controlled for age, sex, race, subjective financial status, highest level of education, and employment status.

### Transparency and Openness

We report in the article how we determined our sample size, all data exclusions, all manipulations, and all measures in the study. This study’s design and analyses were not preregistered. De-identified data, analysis code, and research materials are available from the corresponding author upon reasonable request. All analyses were conducted in R (R Core Team, 2022), and regression models were fit using the *glmmTMB* package (Brooks et al., 2017).

## Results

### Sample Characteristics

Demographic and alcohol-use-related characteristics of the current sample are reported in Table 1. Briefly, participants were an average of 43 years old at baseline, or approximately 50 years old at the time of assessment; 62% were assigned female sex at birth and 80% were white. Participants reported an average of five standard drinks and two drinking days per typical week, and the average AUDIT score in the sample was 3.7. Participants assigned to the adjusting APT did not differ significantly from those assigned to the full APT on any demographic or alcohol-related characteristics (see Table S2), suggesting that our randomization was effective in producing comparable groups.

### Fitting Demand Curves and Estimating Demand Indices from the Adjusting APT

Rates of non-systematic responding were low for both the adjusting and full APTs: 0.46% of participants (*n* = 2) exhibited one or more sign of non-systematic responding on the adjusting APT (reversals from zero: *n* = 1, 0.23%; >10% bounces: *n* = 1, 0.23%; constant or increasing demand: *n* = 2, 0.46%), and 0.65% of participants (*n* = 3) exhibited one or more sign of non-systematic responding on the full APT (reversals from zero: *n* = 1, 0.22%; >10% bounces: *n* = 0, 0%; constant or increasing demand: *n* = 2, 0.43%)^1^. Individual demand curves additionally could not be estimated due to fewer than two points of non-zero consumption for 3.91% of participants (*n* = 17) who completed the adjusting APT, and for 3.25% of participants (*n* = 15) who completed the full APT. Few participants did not reach breakpoint for both the adjusting and full APTs (4.60% and 1.30%, respectively). *R*^2^ values for the individual demand curves estimated from the adjusting APT indicated acceptable fit (*M* = 0.79 ± 0.17, median = 0.82, 8.65% < 0.50) and were comparable to those for the individual demand curves estimated from the full APT (*M* = 0.85 ± 0.10, median = 0.87, 1.13% < 0.50). Plots of the mean aggregate demand curves for both the adjusting and full APTs (Figures 3 and S1) also generally showed acceptable fit of the individual demand curves to the consumption data.

### Relative Efficiency of the Adjusting APT

Participants assigned to the adjusting APT were given an average of 5.91 items (*SD* = 0.28), compared to an average of 11.62 prices (*SD* = 3.53) among those assigned to the full APT, representing an approximate 49% reduction in task length (*t* = 34.64, *df* = 467.28, *p* < 0.001). Similarly, participants spent an average of 1.44 minutes (*SD* = 1.24) completing the adjusting APT compared to an average 1.65 minutes (*SD* = 2.40) completing the full APT, representing an approximate 13% reduction in task duration, though this reduction was not statistically significant (*t* = 1.63, *df* = 697.91, *p* = 0.103).^2^ These reductions in task length and duration account for our use of a full APT with fewer prices than the adjusting APT’s full price set, and for our administration of the full APT in blocks of four to five prices such that participants were not administered price blocks following the first price block wherein they reported any instance of zero consumption. 13.56% of participants who completed the adjusting APT reached breakpoint at prices not included in the price set for the full APT.

### Mean Differences in Demand Indices Between the Adjusting and Full APTs

Descriptive statistics on demand indices estimated from the adjusting and full APTs and mean comparisons between the adjusting and full APTs are reported in Table 2. Mean intensity estimates did not differ significantly between the adjusting and full APTs, both yielding means of approximately four standard drinks when alcohol was free. Mean elasticity estimates likewise did not differ significantly between the adjusting and full APTs. Mean *O*_max_ and breakpoint estimates were significantly higher for the adjusting APT relative to the full APT, with participants reporting an approximate $3 greater maximum expenditure on average and reaching breakpoint approximately $3 higher on average on the adjusting APT compared to the full APT.

**Table 2.** Descriptive statistics on demand indices, overall and stratified by alcohol purchase task version.

|  | Adjusting APT (*n* = 435) |  |  | Full APT (*n* = 462) |  |  | Group comparisons |  |  |  |
| --- | --- | --- | --- | --- | --- | --- | --- | --- | --- | --- |
|  | *M* | *SD* | Med. (Min.–Max.) | *M* | *SD* | Med. (Min.–Max.) | *t* | *df* | *p* | Cohen's *d* |
| Intensity | 3.64 | 2.53 | 3 (0–13) | 3.51 | 2.45 | 3 (0–15) | -0.82 | 883.27 | 0.413 | -0.05 |
| *O*<sub>max</sub> | 22.79 | 18.55 | 22 (0–121) | 20.07 | 13.41 | 18 (0–73) | **-2.5** | **783.49** | **0.013** | **-0.17** |
| Breakpoint | 18.85 | 10.28 | 18 (0–45) | 15.67 | 7.5 | 16 (0–45) | **-5.24** | **787.29** | **<0.001** | **-0.35** |
| Elasticity | 0.009 | 0.008 | 0.006 (0–0.063) | 0.009 | 0.007 | 0.007 (0–0.040) | 0.47 | 837.92 | 0.64 | 0.03 |

*Note.* APT = alcohol purchase task; *M* = mean; *SD* = standard deviation; *df* = degrees of freedom. Bolding denotes statistically significant difference at $\alpha = 0.05$ .

### Convergent Validity of the Adjusting APT

Associations of demand indices from the adjusting and full APTs with alcohol use and problems and differences in these associations between APT versions are summarized in Table 3, with complete results of the multiple regression models examining these associations presented in Table S4. For both the adjusting and full APTs, elevated demand as indicated by all demand indices (greater intensity, *O*_max_, and breakpoint, and lower elasticity) was associated with more standard drinks and drinking days per typical week, a greater likelihood of any HDD in a typical week, and more hazardous drinking. Associations of *O*_max_ with each outcome differed significantly between participants who completed the adjusting versus full APTs, such that associations were stronger for the latter. Associations of intensity, breakpoint, and elasticity with each outcome did not differ significantly between participants who completed the adjusting versus full APTs.

**Table 3.**
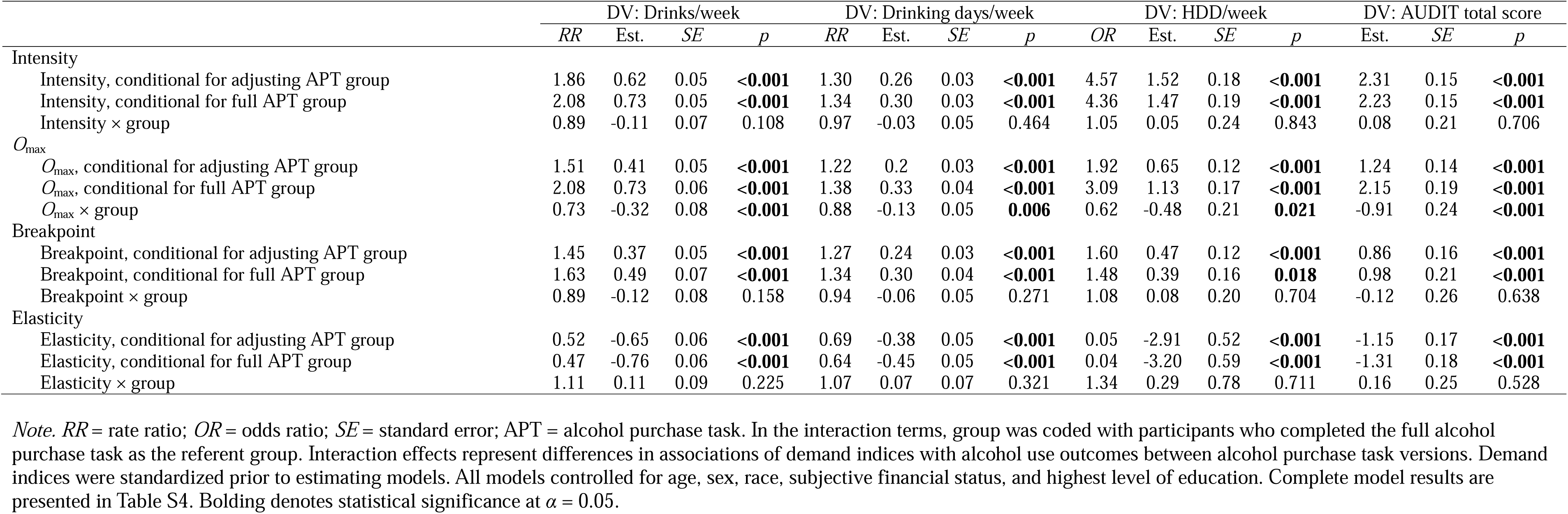
Associations of demand indices with alcohol use and problems for each version of the alcohol purchase task and differences between associations of demand indices with alcohol use and problems between alcohol purchase task versions.

## Discussion

This study aimed to evaluate a brief, idiographic measure of alcohol demand––the adjusting APT––in a large sample of community adults reporting alcohol use. Participants were randomly assigned to complete either the adjusting or full APT, and we compared estimated demand indices and their associations with alcohol use and problems between task versions. Results indicate that the adjusting APT is more efficient than the full APT, reducing task length and duration while still yielding well-fitting demand curves and demand indices that were consistently associated with alcohol use and problems. These findings extend prior validation of the adjusting APT from retrospectively inferred responses among heavy-drinking young adults (Coelho et al., 2026) to stand-alone administration in a broader community sample spanning a wider range of ages and drinking levels, suggesting that the adjusting APT may be a viable option for assessing alcohol demand while minimizing participant burden.

We found that key demand indices could be validly estimated using data from the adjusting APT. Rates of non-systematic responding among participants who completed the adjusting APT were low and comparable to rates among those who completed the full APT, suggesting that the orderliness of the demand data was not compromised by the adjusting APT’s departure from the full APT’s ascending price sequence. The adjusting APT also did not produce greater data loss relative to the full APT from response patterns precluding the estimation of elasticity (i.e., fewer than two prices of non-zero consumption), likely attributable to the adjusting APT’s use of the complete price range from the full APT, administering subsets of items contingent on the individual’s level of alcohol demand. Fit indices and plots of the mean aggregate demand curves further indicated acceptable fit of the individual demand curves estimated from the adjusting APT data, supporting the validity of derived elasticity estimates. This is notable given that the adjusting APT required participants to complete approximately half as many items as the full APT and reduced task completion time. Elasticity is among the demand indices most robustly associated with alcohol use outcomes (Martínez-Loredo et al., 2021; Zvorsky et al., 2019) and may be less susceptible to bias given that it is cognitively implicit and not readily calculable by participants. Thus, the adjusting APT’s ability to validly derive elasticity from just six items has the potential to facilitate robust, multidimensional assessment of alcohol demand across a wider range of research and clinical contexts.

The full and adjusting APTs yielded largely comparable mean levels of alcohol demand, with some exceptions. Mean intensity and elasticity estimates did not differ significantly between participants who completed the two versions of the APT. The similarity of mean intensity levels between the two groups was expected, as intensity corresponds only to the first item of either task. In other words, there was no difference between tasks in the assessment of intensity, and similar mean intensity levels across groups suggests successful randomization. Elasticity, conversely, derives from the full demand curve, estimated from substantially fewer points of demand for the adjusting APT compared to the full APT. That mean elasticity still did not differ significantly between the two groups further supports that the adjusting APT can produce valid demand curves and derived elasticity estimates. By contrast, mean *O*_max_ (i.e., maximum expenditure) and breakpoint estimates were each significantly higher for participants who completed the adjusting APT relative to those who completed the full APT. As participants were randomly assigned to complete either the adjusting or full APT and groups did not differ significantly on any demographic or alcohol-related characteristics, including demand intensity, systematic baseline differences between groups are unlikely to explain these differences in *O*_max_ and breakpoint. These differences are thus more plausibly attributable to differences between the APTs themselves, for example owing to the adjusting APT’s fewer administered prices and thus less information from which to estimate demand indices. Worth noting, however, is that the adjusting APT drew from a more granular set of prices, to which these differences in *O*_max_ and breakpoint between tasks may also be attributable, and thus further research comparing the adjusting APT to a full APT with an identical price set is needed to clarify. Also warranted are future studies comparing demand indices within subjects after administering both the adjusting and full APTs to each participant, as randomization cannot fully rule out that residual group differences on unmeasured characteristics contributed to observed discrepancies in these select demand indices. Nonetheless, differences in *O*_max_ and breakpoint between task versions were relatively small (approximately $3 for each index). Taken together with comparable intensity and elasticity estimates across task versions, our findings support the validity of the adjusting APT as a brief measure of alcohol demand, with potential incremental utility relative to other brief measures, such as the BAAD, that do not permit demand curve estimation or the derivation of elasticity.

Greater alcohol demand as indexed by each demand index from the adjusting APT was associated with more standard drinks and drinking days per typical week, a greater likelihood of any heavy drinking within a typical week, and more hazardous drinking, supporting convergent validity of the adjusting APT. These associations were observed even after adjusting for demographic covariates including age, sex, race, financial status, education level, and employment status. These associations align with those observed between demand indices from the full APT and alcohol use outcomes, both in this study and in prior meta-analytic research (Martínez-Loredo et al., 2021), suggesting that the adjusting APT also captures facets of the relative reinforcing value of alcohol that can explain variability in alcohol use and problems. Notably, in formal comparisons of the direction and magnitude of associations of demand indices with alcohol use outcomes between participants who completed the adjusting and full APTs, only a minority of associations differed significantly between groups. Specifically, associations of *O*_max_ with each alcohol use outcome were weaker among participants who completed the adjusting APT relative to those who completed the full APT, though were still positive and of an expected magnitude among the former group. Thus, the adjusting APT generally appears to preserve explanatory power relative to the full APT, despite its substantially reduced length.

Although our results generally suggest that adjusting APT can efficiently and validly assess alcohol demand, it is worth noting that the adjusting APT is intended to complement, rather than replace, the full APT. For designs requiring high-resolution assessment of demand, such as neuroeconomic functional magnetic resonance imaging studies (e.g., Gray et al., 2017; MacKillop et al., 2014), the full APT may be preferable, as it ultimately provides the most information about participants’ cost-benefit decision making and relative valuation of alcohol. Our data also suggest that despite comprising significantly more items, the full APT did not take participants significantly longer to complete, perhaps owing to the simplicity of estimating consumption across multiple steadily escalating prices and supporting its continued use. Still, the adjusting APT may be favoured when brevity is essential, for example within ecological momentary assessment studies or large clinical batteries, and it appears preferable to the BAAD in these cases given its similar item count yet added ability to produce individual demand curves. The adjusting APT may also be amenable to adaptations to accommodate inflation or typical drink prices in different economic contexts; for example, following a comparable binary-search-style algorithm, the adjusting APT could be adapted to include up to an additional 34 prices while increasing each task permutation length by at most one item. Taken together, investigators should select the APT best suited for their study design and research questions, appreciating the caveat with the adjusting APT that although comparable, its resultant demand indices should not be presumed identical to those that would have been obtained from a full APT.

## Limitations

Several limitations of this study should be considered. First, the adjusting and full APTs were respectively administered to two independent groups, and thus task comparisons on demand index estimates and associations of demand indices with alcohol use outcomes are between groups that may differ on unmeasured characteristics, rather than within subjects. Though randomization to the adjusting versus full APT was used to mitigate systematic differences between the groups, within-subjects designs are needed to further validate the adjusting APT in comparison to the full APT. In addition, the adjusting APT assesses hypothetical alcohol consumption, which may deviate from actual alcohol consumption. Standard APTs have been shown to perform similarly for hypothetical and actual rewards (Amlung et al., 2012), but whether this extends to the adjusting APT has not been empirically tested. Further, the majority of participants in our sample were White, cisgender, financially stable, and had achieved a high level of education, potentially limiting generalizability to the broader population. As this was a community sample, we also had low representation of hazardous drinking (and presumably alcohol use disorder), and thus further validation of the adjusting APT in heavy-drinking and clinical samples is needed.

## Conclusions

In summary, this study provides support for the validity of the adjusting APT as a brief, multidimensional measure of behavioural economic demand for alcohol. Findings suggest that this measure’s idiographic approach to assessing alcohol demand reduced participant burden relative to a fixed-item APT, yet still produced valid estimates of demand indices that were associated with key alcohol use outcomes. Inflation of estimates was present for *O*_max_ and breakpoint, but was modest in magnitude. Nonetheless, the adjusting APT should not be assumed to produce identical estimates to the traditional APT for these indices. Future research should further evaluate the psychometric properties of the adjusting APT, including using within-subjects designs wherein participants each complete both the adjusting APT and its full-length counterpart. Other future research directions include evaluating the adjusting APT’s performance as a measure of intraindividual changes in alcohol demand across experimental and naturalistic settings.

## Data Availability

De-identified data, analysis code, and research materials are available from the corresponding author upon reasonable request.

## Footnotes

1 In a sensitivity analysis applying a more conservative bounce criterion of bounces at any (rather than more than 10% of) price increments, *n* = 1 (0.23%) exhibited one or more bounces on the adjusting APT and *n* = 4 (0.87%) exhibited one or more bounces on the full APT.

2 Extreme outliers in completion time, defined as completion times more than 3.99 standard deviations from the mean or that were visibly disjointed from the distribution for a given APT version, were removed prior to calculating descriptive statistics, as these presumably represented cases when participants stepped away from the questionnaire part way through completion.

## References

Acker, J., & MacKillop, J. (2013). Behavioral Economic Analysis of Cue-Elicited Craving for Tobacco: A Virtual Reality Study. Nicotine & Tobacco Research, 15(8), 1409–1416. 10.1093/ntr/nts341

Acuff, S. F., Amlung, M., Dennhardt, A. A., MacKillop, J., & Murphy, J. G. (2020). Experimental manipulations of behavioral economic demand for addictive commodities: A meta-analysis. *Addiction (Abingdon*, England), 115(5), 817–831. 10.1111/add.14865

Acuff, S. F., MacKillop, J., & Murphy, J. G. (2023). A contextualized reinforcer pathology approach to addiction. Nature Reviews Psychology, 2(5), Article 5. 10.1038/s44159-023-00167-y

Amlung, M. T., Acker, J., Stojek, M. K., Murphy, J. G., & MacKillop, J. (2012). Is talk “cheap”? An initial investigation of the equivalence of alcohol purchase task performance for hypothetical and actual rewards. *Alcoholism*, Clinical and Experimental Research, 36(4), 716–724. 10.1111/j.1530-0277.2011.01656.x

Amlung, M. T., Marsden, E., Hargreaves, T., Sweet, L. H., Murphy, J. G., & MacKillop, J. (2024). Neural correlates of increased alcohol demand following alcohol cue exposure in adult heavy drinkers. Psychiatry Research: Neuroimaging, 340, 111809. 10.1016/j.pscychresns.2024.111809

Amlung, M. T., McCarty, K. N., Morris, D. H., Tsai, C.-L., & McCarthy, D. M. (2015). Increased behavioral economic demand and craving for alcohol following a laboratory alcohol challenge. *Addiction (Abingdon*, England), 110(9), 1421–1428. 10.1111/add.12897

Aston, E. R., & Merrill, J. E. (2023). Alcohol demand assessed daily as a predictor of same day drinking. Psychology of Addictive Behaviors: Journal of the Society of Psychologists in Addictive Behaviors, 37(1), 114–120. 10.1037/adb0000890

Bickel, W. K., Johnson, M. W., Koffarnus, M. N., MacKillop, J., & Murphy, J. G. (2014). The behavioral economics of substance use disorders: Reinforcement pathologies and their repair. Annual Review of Clinical Psychology, 10, 641–677. 10.1146/annurev-clinpsy-032813-153724

Bird, B. M., Belisario, K., Minhas, M., Acuff, S. F., Ferro, M. A., Amlung, M. T., Murphy, J. G., & MacKillop, J. (2024). Longitudinal examination of alcohol demand and alcohol-related reinforcement as predictors of heavy drinking and adverse alcohol consequences in emerging adults. Addiction, n/a. 10.1111/add.16443

Brooks, M. E., Kristensen, K., Benthem, K. J. van, Magnusson, A., Berg, C. W., Nielsen, A., Skaug, H. J., Mächler, M., & Bolker, B. M. (2017). glmmTMB Balances Speed and Flexibility Among Packages for Zero-inflated Generalized Linear Mixed Modeling. The R Journal, 9(2), 378–400.

Cassidy, R. N., Bernstein, M. H., Magill, M., MacKillop, J., Murphy, J. G., & Colby, S. M. (2019). Alcohol demand moderates brief motivational intervention outcomes in underage young adult drinkers. Addictive Behaviors, 98, 106044. 10.1016/j.addbeh.2019.106044

Coelho, S. G., Belisario, K. L., Keough, M. T., Amlung, M. T., Murphy, J. G., & MacKillop, J. (2026). Development of a brief adjusting alcohol purchase task as a measure of behavioral economic demand for alcohol in two samples of heavy-drinking young adults. Psychological Assessment, 38(3), 203–219. 10.1037/pas0001434

Collins, R. L., Parks, G. A., & Marlatt, G. A. (1985). Social determinants of alcohol consumption: The effects of social interaction and model status on the self-administration of alcohol. Journal of Consulting and Clinical Psychology, 53(2), 189–200. 10.1037//0022-006x.53.2.189

Gray, J. C., Amlung, M. T., Owens, M., Acker, J., Brown, C. L., Brody, G. H., Sweet, L. H., & MacKillop, J. (2017). The Neuroeconomics of Tobacco Demand: An Initial Investigation of the Neural Correlates of Cigarette Cost-Benefit Decision Making in Male Smokers. Scientific Reports, 7(1), 41930. 10.1038/srep41930

Hardy, L., Bakou, A. E., Shuai, R., Acuff, S. F., MacKillop, J., Murphy, C. M., Murphy, J. G., & Hogarth, L. (2021). Associations between the Brief Assessment of Alcohol Demand (BAAD) questionnaire and alcohol use disorder severity in UK samples of student and community drinkers. Addictive Behaviors, 113, 106724. 10.1016/j.addbeh.2020.106724

Harris, P. A., Taylor, R., Minor, B. L., Elliott, V., Fernandez, M., O’Neal, L., McLeod, L., Delacqua, G., Delacqua, F., Kirby, J., Duda, S. N., & REDCap Consortium. (2019). The REDCap consortium: Building an international community of software platform partners. Journal of Biomedical Informatics, 95, 103208. 10.1016/j.jbi.2019.103208

Hursh, S. R. (1980). Economic Concepts for the Analysis of Behavior. Journal of the Experimental Analysis of Behavior, 34(2), 219–238. 10.1901/jeab.1980.34-219

Hursh, S. R., Galuska, C. M., Winger, G., & Woods, J. H. (2005). The economics of drug abuse: A quantitative assessment of drug demand. Molecular Interventions, 5(1), 20–28. 10.1124/mi.5.1.6

Hursh, S. R., & Silberberg, A. (2008). Economic demand and essential value. Psychological Review, 115(1), 186–198. 10.1037/0033-295X.115.1.186

Kaplan, B. A., Gilroy, S. P., Reed, D. D., Koffarnus, M. N., & Hursh, S. R. (2019). The R package beezdemand: Behavioral Economic Easy Demand. Perspectives on Behavior Science, 42(1), 163–180. 10.1007/s40614-018-00187-7

Koffarnus, M. N., Franck, C. T., Stein, J. S., & Bickel, W. K. (2015). A modified exponential behavioral economic demand model to better describe consumption data. Experimental and Clinical Psychopharmacology, 23, 504–512. 10.1037/pha0000045

MacKillop, J. (2016). The Behavioral Economics and Neuroeconomics of Alcohol Use Disorders. *Alcoholism*, Clinical and Experimental Research, 40(4), 672–685. 10.1111/acer.13004

MacKillop, J., Amlung, M. T., Acker, J., Gray, J. C., Brown, C. L., Murphy, J. G., Ray, L. A., & Sweet, L. H. (2014). The Neuroeconomics of Alcohol Demand: An Initial Investigation of the Neural Correlates of Alcohol Cost–Benefit Decision Making in Heavy Drinking Men. Neuropsychopharmacology, 39(8), 1988–1995. 10.1038/npp.2014.47

MacKillop, J., & Murphy, J. G. (2007). A behavioral economic measure of demand for alcohol predicts brief intervention outcomes. Drug and Alcohol Dependence, 89, 227–233. 10.1016/j.drugalcdep.2007.01.002

Martínez-Loredo, V., González-Roz, A., Secades-Villa, R., Fernández-Hermida, J. R., & MacKillop, J. (2021). Concurrent validity of the Alcohol Purchase Task for measuring the reinforcing efficacy of alcohol: An updated systematic review and meta-analysis. Addiction, 116(10), 2635–2650. 10.1111/add.15379

Merrill, J. E., & Aston, E. R. (2020). Alcohol demand assessed daily: Validity, variability, and the influence of drinking-related consequences. Drug and Alcohol Dependence, 208, 107838. 10.1016/j.drugalcdep.2020.107838

Murphy, J. G., Dennhardt, A. A., Yurasek, A. M., Skidmore, J. R., Martens, M. P., MacKillop, J., & McDevitt-Murphy, M. E. (2015). Behavioral economic predictors of brief alcohol intervention outcomes. Journal of Consulting and Clinical Psychology, 83(6), 1033–1043. 10.1037/ccp0000032

Murphy, J. G., & MacKillop, J. (2006). Relative reinforcing efficacy of alcohol among college student drinkers. Experimental and Clinical Psychopharmacology, 14, 219–227. 10.1037/1064-1297.14.2.219

Owens, M. M., Murphy, C. M., & MacKillop, J. (2015). Initial Development of a Brief Behavioral Economic Assessment of Alcohol Demand. *Psychology of Consciousness (Washington*, D.C.), 2(2), 144–152. 10.1037/cns0000056

R Core Team. (2022). R: A language and environment for statistical computing [Computer software]. R Foundation for Statistical Computing. https://www.R-project.org/

Saunders, J. B., Aasland, O. G., Babor, T. F., de la Fuente, J. R., & Grant, M. (1993). Development of the Alcohol Use Disorders Identification Test (AUDIT): WHO Collaborative Project on Early Detection of Persons with Harmful Alcohol Consumption--II. *Addiction (Abingdon*, England), 88(6), 791–804. 10.1111/j.1360-0443.1993.tb02093.x

Stein, J. S., Koffarnus, M. N., Snider, S. E., Quisenberry, A. J., & Bickel, W. K. (2015). Identification and Management of Nonsystematic Purchase-Task Data: Towards Best Practice. Experimental and Clinical Psychopharmacology, 23(5), 377–386. 10.1037/pha0000020

Zvorsky, I., Nighbor, T. D., Kurti, A. N., DeSarno, M., Naudé, G., Reed, D. D., & Higgins, S. T. (2019). Sensitivity of hypothetical purchase task indices when studying substance use: A systematic literature review. Preventive Medicine, 128, 105789. 10.1016/j.ypmed.2019.105789

