## Supplemental Materials for "Evaluating an Adjusting Alcohol Purchase Task as a Brief Measure of Behavioural Economic Demand for Alcohol in a Large Sample of Community Adults"

**Alcohol Purchase Task Instructional Set**

Instructions for the Alcohol Purchase Task (both full-length and adjusting) were as follows: “Please respond to these questions as if you were actually in a TYPICAL SITUATION when you drink alcohol. Imagine where you typically drink, what you typically drink, and who you typically drink with, if anyone. The available drinks are standard size beer (12 oz.), wine (5 oz.), shots of hard liquor (1.5 oz.), or mixed drinks containing one shot of liquor. Assume that you did not drink alcohol before you are making these decisions, and will not have an opportunity to drink elsewhere after making these decisions. In addition, assume that you would consume every drink you request; that is, you cannot stockpile drinks for a later date or bring drinks home with you.”

For the full alcohol purchase task, participants were presented with 30 different prices, administered in the following order: $0, $0.25, $0.5, $1, $1.5, $2, $2.5, $3, $4, $5, $6, $7, $8, $9, $10, $11, $12, $13, $14, $15, $16, $18, $20, $22, $24, $26, $28, $30, $35, $40).

For the adjusting alcohol purchase task, participants were presented with one of the following sequences of prices, according to the adjusting alcohol purchase task algorithm (see Figure 1):

1. $0, $11, $22, $30, $40

2. $0, $11, $22, $30, $40, $35

3. $0, $11, $22, $30, $26, $28

4. $0, $11, $22, $30, $26, $24

5. $0, $11, $22, $15, $18, $20

6. $0, $11, $22, $15, $18, $16

7. $0, $11, $22, $15, $13, $14

8. $0, $11, $22, $15, $13, $12

9. $0, $11, $3, $7, $9, $10

10. $0, $11, $3, $7, $9, $8

11. $0, $11, $3, $7, $5, $6

12. $0, $11, $3, $7, $5, $4

13. $0, $11, $3, $1, $2, $2.50

14. $0, $11, $3, $1, $2, $1.50

15. $0, $11, $3, $1, $0.25, $0.50

16. $0, $11, $3, $1, $0.25

**Table S1. Characteristics of participants from the larger cohort study who were included in versus excluded from the current study sample**

|  |  | Included in study sample  (*n* = 897) | | Excluded from study sample  (*n* = 605) | | Group comparisons | | |
| --- | --- | --- | --- | --- | --- | --- | --- | --- |
|  |  | *M* or *n* | *SD* or % | *M* or *n* | *SD* or % | *t* or *X*^2^ | *df* | *p* |
| Baseline age (years) | | 35.79 | 14.45 | 32.8 | 12.93 | **4.19** | **1387** | **< 0.001** |
| Sex: female (vs. male) | | 553 | 61.65 | 343 | 56.69 | 3.48 | 1 | 0.062 |
| Race: white (vs. non-white) | | 723 | 80.60 | 470 | 77.69 | 1.71 | 1 | 0.192 |
| Highest level of education: ≥ bachelor's degree (vs. < bachelor's degree) | | 428 | 47.71 | 251 | 41.49 | **5.41** | **1** | **0.020** |
| Employment status: employed (vs. not employed) | | 705 | 78.60 | 459 | 75.87 | 1.39 | 1 | 0.239 |
| Subjective financial status | |  |  |  |  | **15.20** | **3** | **0.002** |
|  | Not enough to pay some bills no matter how hard you try | 23 | 2.56 | 28 | 4.63 |  |  |  |
|  | Enough to pay bills, but have had to cut back | 162 | 18.06 | 141 | 23.31 |  |  |  |
|  | Enough to pay bills without cutting back but no “extras” | 283 | 31.55 | 197 | 32.56 |  |  |  |
|  | Enough money for “extras” | **429** | **47.83** | **239** | **39.50** |  |  |  |

*Note. M* = mean; *SD* = standard deviation; *df* = degrees of freedom. Bolding denotes statistically significant difference at *α* = 0.05.

**Table S2. Sample characteristics, stratified by alcohol purchase task version**

|  |  | Adjusting APT  (*n* = 435) | | Full APT  (*n* = 462) | | Group comparisons | | |
| --- | --- | --- | --- | --- | --- | --- | --- | --- |
|  |  | *n* | % | *n* | % | *X*^2^ | *df* | *p* |
| Sex | |  |  |  |  | <0.01 | 1.00 | 0.965 |
|  | Male | 166 | 38.16 | 178 | 38.53 |  |  |  |
|  | Female | 269 | 61.84 | 284 | 61.47 |  |  |  |
| Gender | |  |  |  |  | 1.96 | 2.00 | 0.376 |
|  | Man | 165 | 37.93 | 175 | 37.88 |  |  |  |
|  | Woman | 265 | 60.92 | 276 | 59.74 |  |  |  |
|  | Transgender/nonbinary | 5 | 1.15 | 11 | 2.38 |  |  |  |
| Race | |  |  |  |  | 4.14 | 9.00 | 0.902 |
|  | Multiple | 18 | 4.14 | 18 | 3.90 |  |  |  |
|  | White | 350 | 80.46 | 365 | 79.00 |  |  |  |
|  | Black | 7 | 1.61 | 9 | 1.95 |  |  |  |
|  | East Asian | 17 | 3.91 | 21 | 4.55 |  |  |  |
|  | South Asian | 18 | 4.14 | 15 | 3.25 |  |  |  |
|  | Southeast Asian | 4 | 0.92 | 2 | 0.43 |  |  |  |
|  | Middle Eastern | 6 | 1.38 | 7 | 1.52 |  |  |  |
|  | First Nations/Indigenous | 3 | 0.69 | 3 | 0.65 |  |  |  |
|  | Pacific Islander | 1 | 0.23 | 2 | 0.43 |  |  |  |
|  | Other | 11 | 2.53 | 20 | 4.33 |  |  |  |
| Latinx | |  |  |  |  | <0.01 | 1.00 | >0.999 |
|  | No | 418 | 96.09 | 444 | 96.10 |  |  |  |
|  | Yes | 17 | 3.91 | 18 | 3.90 |  |  |  |
| Subjective financial status | |  |  |  |  | 1.39 | 3.00 | 0.707 |
|  | Not enough to pay some bills no matter how hard you try | 19 | 4.37 | 16 | 3.46 |  |  |  |
|  | Enough to pay bills, but have had to cut back | 97 | 22.30 | 96 | 20.78 |  |  |  |
|  | Enough to pay bills without cutting back but no “extras” | 115 | 26.44 | 117 | 25.32 |  |  |  |
|  | Enough money for “extras” | 204 | 46.90 | 233 | 50.43 |  |  |  |
| Annual household income | |  |  |  |  | 13.64 | 10.00 | 0.190 |
|  | < $15,000 | 13 | 2.99 | 8 | 1.73 |  |  |  |
|  | $15,000 to < $30,000 | 14 | 3.22 | 19 | 4.11 |  |  |  |
|  | $30,000 to < $45,000 | 20 | 4.60 | 33 | 7.14 |  |  |  |
|  | $45,000 to < $60,000 | 27 | 6.21 | 32 | 6.93 |  |  |  |
|  | $60,000 to < $75,000 | 42 | 9.66 | 36 | 7.79 |  |  |  |
|  | $79,000 to < $90,000 | 38 | 8.74 | 46 | 9.96 |  |  |  |
|  | $90,000 to < $105,000 | 57 | 13.10 | 36 | 7.79 |  |  |  |
|  | $105,000 to < $120,000 | 47 | 10.80 | 47 | 10.17 |  |  |  |
|  | $120,000 to < $135,000 | 26 | 5.98 | 25 | 5.41 |  |  |  |
|  | $135,000 to < $150,000 | 29 | 6.67 | 30 | 6.49 |  |  |  |
|  | ≥ $150,000 | 122 | 28.05 | 150 | 32.47 |  |  |  |
| Highest level of education | |  |  |  |  | 6.17 | 5.00 | 0.290 |
|  | High school graduate (or GED) or less | 24 | 5.52 | 23 | 4.98 |  |  |  |
|  | Some college/university | 112 | 25.75 | 100 | 21.65 |  |  |  |
|  | Associates degree | 43 | 9.89 | 42 | 9.09 |  |  |  |
|  | Bachelor’s degree | 134 | 30.80 | 175 | 37.88 |  |  |  |
|  | Master’s degree | 75 | 17.24 | 68 | 14.72 |  |  |  |
|  | Professional degree | 47 | 10.80 | 54 | 11.69 |  |  |  |
| Employment Status | |  |  |  |  | 3.04 | 2.00 | 0.218 |
|  | Employed full-time | 283 | 65.06 | 309 | 66.88 |  |  |  |
|  | Employed part-time | 54 | 12.41 | 41 | 8.87 |  |  |  |
|  | Not employed | 98 | 22.53 | 112 | 24.24 |  |  |  |
|  |  | *M* | *SD* | *M* | *SD* | *t* | *df* | *p* |
| Baseline age | | 42.70 | 14.24 | 43.01 | 14.68 | 0.32 | 894.17 | 0.750 |
| Drinks/week | | 4.66 | 5.62 | 4.77 | 6.40 | 0.27 | 890.74 | 0.791 |
| Drinking days/week | | 2.21 | 1.78 | 2.20 | 1.83 | -0.03 | 893.91 | 0.977 |
| HDD/week | | 0.25 | 0.85 | 0.34 | 1.14 | 1.27 | 852.22 | 0.204 |
| AUDIT total score | | 3.79 | 3.73 | 3.69 | 3.84 | -0.41 | 894.15 | 0.685 |

*Note.* APT = alcohol purchase task; *M* = mean; *SD* = standard deviation; *df* = degrees of freedom; HDD = heavy drinking days; AUDIT = Alcohol Use Disorder Identification Test.

**Table S3. Frequencies of task permutations among participants randomized to the adjusting alcohol purchase task**

| Adjusting APT permutation | *n* | % |
| --- | --- | --- |
| 1. $0, $11, $22, $30, $40 | 17 | 3.91 |
| 2. $0, $11, $22, $30, $40, $35 | 2 | 0.46 |
| 3. $0, $11, $22, $30, $26, $28 | 2 | 0.46 |
| 4. $0, $11, $22, $30, $26, $24 | 3 | 0.69 |
| 5. $0, $11, $22, $15, $18, $20 | 5 | 1.15 |
| 6. $0, $11, $22, $15, $18, $16 | 15 | 3.45 |
| 7. $0, $11, $22, $15, $13, $14 | 30 | 6.9 |
| 8. $0, $11, $22, $15, $13, $12 | 13 | 2.99 |
| 9. $0, $11, $3, $7, $9, $10 | 55 | 12.64 |
| 10. $0, $11, $3, $7, $9, $8 | 31 | 7.13 |
| 11. $0, $11, $3, $7, $5, $6 | 72 | 16.55 |
| 12. $0, $11, $3, $7, $5, $4 | 33 | 7.59 |
| 13. $0, $11, $3, $1, $2, $2.50 | 88 | 20.23 |
| 14. $0, $11, $3, $1, $2, $1.50 | 17 | 3.91 |
| 15. $0, $11, $3, $1, $0.25, $0.50 | 31 | 7.13 |
| 16. $0, $11, $3, $1, $0.25 | 21 | 4.83 |

*Note.* APT = alcohol purchase task.

**Table S4. Results of regression models examining associations of demand indices with alcohol use and problems and differences in these associations by alcohol purchase task version**

|  |  | DV: Drinks/week | | | | DV: Drinking days/week | | | | DV: HDD/week | | | | DV: AUDIT total score | | |
| --- | --- | --- | --- | --- | --- | --- | --- | --- | --- | --- | --- | --- | --- | --- | --- | --- |
|  |  | *RR* | Est. | *SE* | *p* | *RR* | Est. | *SE* | *p* | *OR* | Est. | *SE* | *p* | Est. | *SE* | *p* |
| Intensity | |  |  |  |  |  |  |  |  |  |  |  |  |  |  |  |
|  | Intensity | 2.08 | 0.73 | 0.05 | **<0.001** | 1.34 | 0.30 | 0.03 | **<0.001** | 4.36 | 1.47 | 0.19 | **<0.001** | 2.23 | 0.15 | **<0.001** |
|  | Group | 1.01 | 0.01 | 0.06 | 0.849 | 1.00 | 0.00 | 0.05 | 0.924 | 0.83 | -0.19 | 0.30 | 0.523 | -0.02 | 0.21 | 0.916 |
|  | Age | 1.02 | 0.02 | 0.00 | **<0.001** | 1.01 | 0.01 | 0.00 | **<0.001** | 1.01 | 0.01 | 0.01 | 0.536 | 0.01 | 0.01 | 0.198 |
|  | Sex | 0.95 | -0.05 | 0.07 | 0.413 | 0.98 | -0.02 | 0.05 | 0.672 | 2.36 | 0.86 | 0.28 | **0.002** | -0.04 | 0.22 | 0.847 |
|  | Race | 1.15 | 0.14 | 0.08 | 0.096 | 1.17 | 0.15 | 0.07 | **0.026** | 1.19 | 0.18 | 0.34 | 0.605 | 0.23 | 0.27 | 0.382 |
|  | Subjective financial status | 1.06 | 0.06 | 0.04 | 0.107 | 1.06 | 0.06 | 0.03 | **0.034** | 1.04 | 0.04 | 0.15 | 0.797 | 0.14 | 0.12 | 0.259 |
|  | Highest level of education | 0.98 | -0.02 | 0.07 | 0.787 | 1.04 | 0.04 | 0.06 | 0.524 | 0.72 | -0.33 | 0.28 | 0.237 | -0.16 | 0.23 | 0.482 |
|  | Employment status | 0.95 | -0.05 | 0.09 | 0.554 | 0.94 | -0.06 | 0.07 | 0.402 | 1.04 | 0.04 | 0.39 | 0.915 | -0.34 | 0.29 | 0.250 |
|  | Intensity × group | 0.89 | -0.11 | 0.07 | 0.108 | 0.97 | -0.03 | 0.05 | 0.464 | 1.05 | 0.05 | 0.24 | 0.843 | 0.08 | 0.21 | 0.706 |
| *O*_max_ | |  |  |  |  |  |  |  |  |  |  |  |  |  |  |  |
|  | *O*_max_ | 2.08 | 0.73 | 0.06 | **<0.001** | 1.38 | 0.33 | 0.04 | **<0.001** | 3.09 | 1.13 | 0.17 | **<0.001** | 2.15 | 0.19 | **<0.001** |
|  | Group | 0.99 | -0.01 | 0.07 | 0.871 | 0.97 | -0.03 | 0.05 | 0.557 | 1.01 | 0.01 | 0.24 | 0.970 | -0.21 | 0.23 | 0.352 |
|  | Age | 1.00 | 0.00 | 0.00 | 0.131 | 1.01 | 0.01 | 0.00 | **<0.001** | 0.97 | -0.03 | 0.01 | **0.017** | -0.02 | 0.01 | 0.081 |
|  | Sex | 0.75 | -0.28 | 0.07 | **<0.001** | 0.89 | -0.12 | 0.05 | **0.019** | 1.08 | 0.08 | 0.23 | 0.743 | -0.82 | 0.23 | **<0.001** |
|  | Race | 1.21 | 0.19 | 0.09 | **0.037** | 1.19 | 0.18 | 0.07 | **0.010** | 1.25 | 0.22 | 0.30 | 0.460 | 0.30 | 0.29 | 0.310 |
|  | Subjective financial status | 0.98 | -0.02 | 0.04 | 0.631 | 1.04 | 0.04 | 0.03 | 0.204 | 0.93 | -0.07 | 0.13 | 0.570 | -0.01 | 0.13 | 0.939 |
|  | Highest level of education | 0.87 | -0.14 | 0.07 | 0.060 | 1.00 | 0.00 | 0.06 | 0.967 | 0.56 | -0.57 | 0.25 | **0.022** | -0.51 | 0.25 | **0.042** |
|  | Employment status | 0.84 | -0.17 | 0.10 | 0.077 | 0.89 | -0.11 | 0.07 | 0.117 | 0.85 | -0.16 | 0.34 | 0.643 | -0.55 | 0.32 | 0.088 |
|  | *O*_max_ × group | 0.73 | -0.32 | 0.08 | **<0.001** | 0.88 | -0.13 | 0.05 | **0.006** | 0.62 | -0.48 | 0.21 | **0.021** | -0.91 | 0.24 | **<0.001** |
| Breakpoint | |  |  |  |  |  |  |  |  |  |  |  |  |  |  |  |
|  | Breakpoint | 1.63 | 0.49 | 0.07 | **<0.001** | 1.34 | 0.30 | 0.04 | **<0.001** | 1.48 | 0.39 | 0.16 | **0.018** | 0.98 | 0.21 | **<0.001** |
|  | Group | 0.86 | -0.15 | 0.07 | **0.040** | 0.91 | -0.10 | 0.05 | 0.060 | 0.81 | -0.21 | 0.22 | 0.350 | -0.27 | 0.25 | 0.277 |
|  | Age | 1.00 | 0.00 | 0.00 | 0.817 | 1.01 | 0.01 | 0.00 | **0.001** | 0.97 | -0.03 | 0.01 | **0.003** | -0.03 | 0.01 | **0.010** |
|  | Sex | 0.68 | -0.39 | 0.07 | **<0.001** | 0.83 | -0.19 | 0.05 | **<0.001** | 0.90 | -0.11 | 0.22 | 0.613 | -1.07 | 0.25 | **<0.001** |
|  | Race | 1.25 | 0.23 | 0.09 | **0.016** | 1.19 | 0.17 | 0.07 | **0.013** | 1.17 | 0.15 | 0.27 | 0.575 | 0.33 | 0.31 | 0.295 |
|  | Subjective financial status | 0.99 | -0.01 | 0.04 | 0.849 | 1.03 | 0.03 | 0.03 | 0.323 | 0.98 | -0.03 | 0.12 | 0.838 | 0.03 | 0.14 | 0.848 |
|  | Highest level of education | 0.73 | -0.32 | 0.08 | **<0.001** | 0.93 | -0.08 | 0.06 | 0.169 | 0.46 | -0.77 | 0.23 | **0.001** | -0.84 | 0.27 | **0.002** |
|  | Employment status | 0.81 | -0.21 | 0.10 | **0.045** | 0.87 | -0.14 | 0.07 | 0.054 | 0.89 | -0.11 | 0.32 | 0.716 | -0.47 | 0.35 | 0.182 |
|  | Breakpoint × group | 0.89 | -0.12 | 0.08 | 0.158 | 0.94 | -0.06 | 0.05 | 0.271 | 1.08 | 0.08 | 0.20 | 0.704 | -0.12 | 0.26 | 0.638 |
| Elasticity | |  |  |  |  |  |  |  |  |  |  |  |  |  |  |  |
|  | Elasticity | 0.47 | -0.76 | 0.06 | **<0.001** | 0.64 | -0.45 | 0.05 | **<0.001** | 0.04 | -3.20 | 0.59 | **<0.001** | -1.31 | 0.18 | **<0.001** |
|  | Group | 1.03 | 0.03 | 0.07 | 0.620 | 1.01 | 0.01 | 0.05 | 0.879 | 1.10 | 0.09 | 0.51 | 0.853 | 0.03 | 0.25 | 0.894 |
|  | Age | 1.01 | 0.01 | 0.00 | **0.034** | 1.01 | 0.01 | 0.00 | **<0.001** | 0.98 | -0.02 | 0.01 | 0.117 | -0.02 | 0.01 | 0.170 |
|  | Sex | 0.77 | -0.26 | 0.07 | **<0.001** | 0.90 | -0.11 | 0.05 | **0.026** | 1.01 | 0.01 | 0.23 | 0.953 | -0.79 | 0.25 | **0.002** |
|  | Race | 1.15 | 0.14 | 0.09 | 0.096 | 1.13 | 0.12 | 0.07 | 0.060 | 1.05 | 0.05 | 0.29 | 0.871 | 0.29 | 0.32 | 0.370 |
|  | Subjective financial status | 1.01 | 0.01 | 0.04 | 0.884 | 1.04 | 0.04 | 0.03 | 0.199 | 0.91 | -0.10 | 0.13 | 0.449 | 0.03 | 0.14 | 0.831 |
|  | Highest level of education | 0.77 | -0.26 | 0.07 | **<0.001** | 0.93 | -0.07 | 0.05 | 0.178 | 0.48 | -0.74 | 0.25 | **0.003** | -0.84 | 0.27 | **0.002** |
|  | Employment status | 0.85 | -0.17 | 0.10 | 0.078 | 0.87 | -0.14 | 0.07 | **0.035** | 0.71 | -0.34 | 0.35 | 0.340 | -0.48 | 0.36 | 0.179 |
|  | Elasticity × group | 1.11 | 0.11 | 0.09 | 0.225 | 1.07 | 0.07 | 0.07 | 0.321 | 1.34 | 0.29 | 0.78 | 0.711 | 0.16 | 0.25 | 0.528 |

*Note. RR* = rate ratio; *OR* = odds ratio; *SE* = standard error; APT = alcohol purchase task. Group was coded as 0 = full alcohol purchase task, 1 = adjusting alcohol purchase task; age was coded in years; sex was coded as 0 = male, 1 = female; race was coded as 0 = non-white, 1 = white; highest level of education was coded as 0 = less than bachelor’s degree, 1 = bachelor’s degree or higher; and employment status was coded as 0 = not employed, 1 = employed. Interaction effects represent differences in associations of demand indices with alcohol use outcomes between alcohol purchase task versions. Demand indices were standardized prior to estimating models. Bolding denotes statistical significance at *α* = 0.05.

**Figure S1. Mean Aggregate Demand Curves for the Full Alcohol Purchase Task and Each Permutation of the Adjusting Alcohol Purchase Task**

| **Full APT (*n* = 444)**  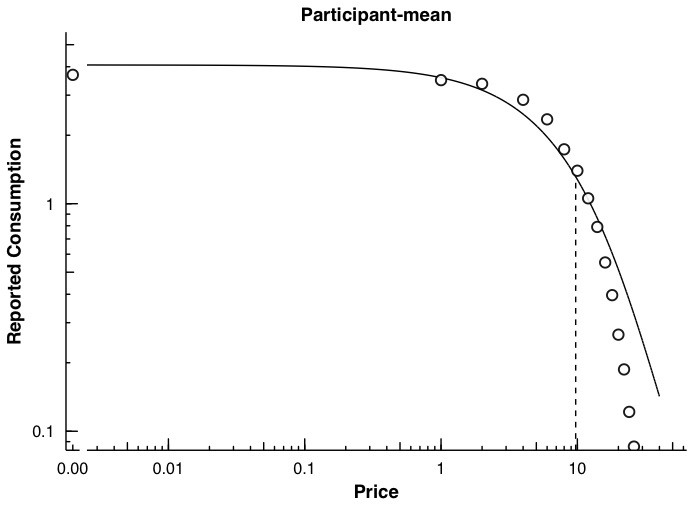 | **Adjusting APT Permutation 2 (*n* = 1)**  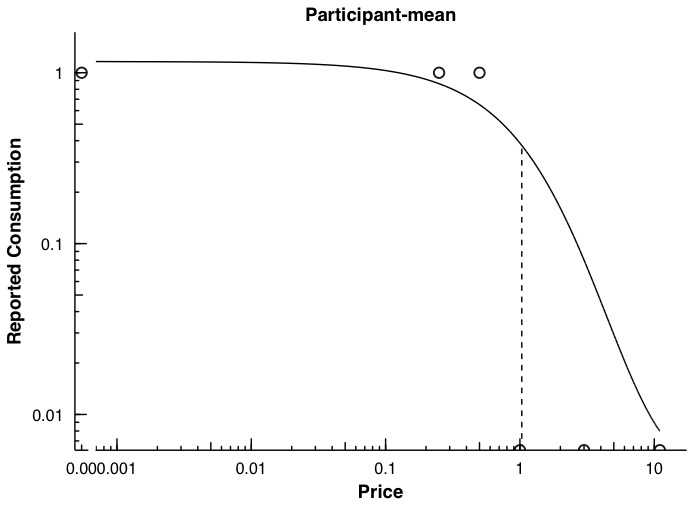 |
| --- | --- |
| **Adjusting APT Permutation 3 (*n* = 2)**  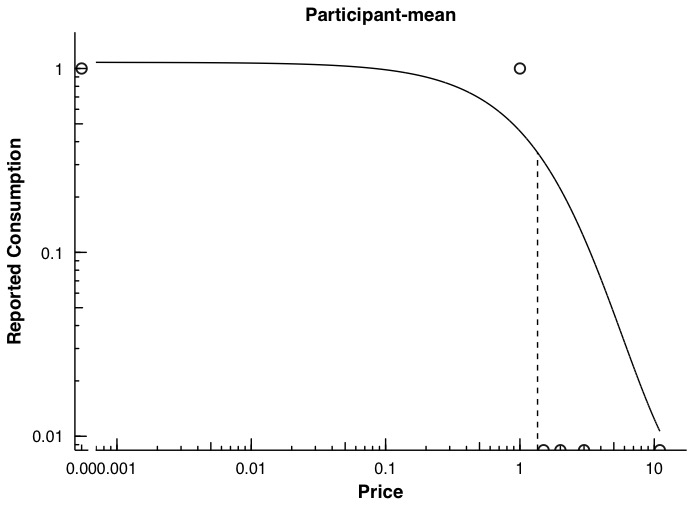 | **Adjusting APT Permutation 4 (*n* = 3)**  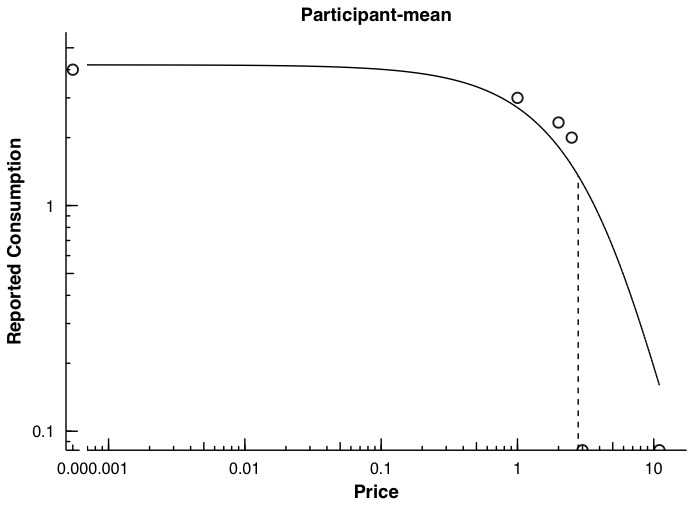 |
| **Adjusting APT Permutation 5 (*n* = 5)**  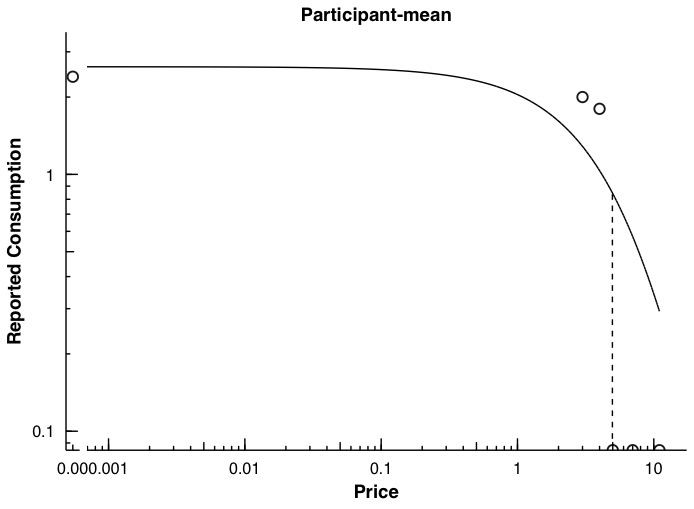 | **Adjusting APT Permutation 6 (*n* = 15)**  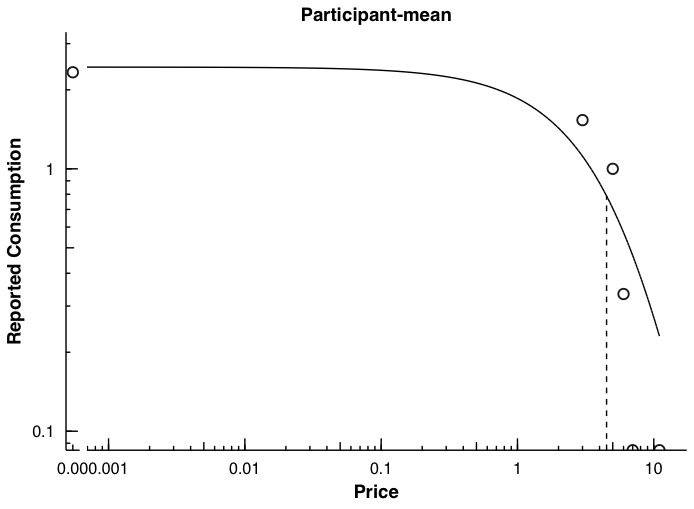 |
| **Adjusting APT Permutation 7 (*n* = 30)**  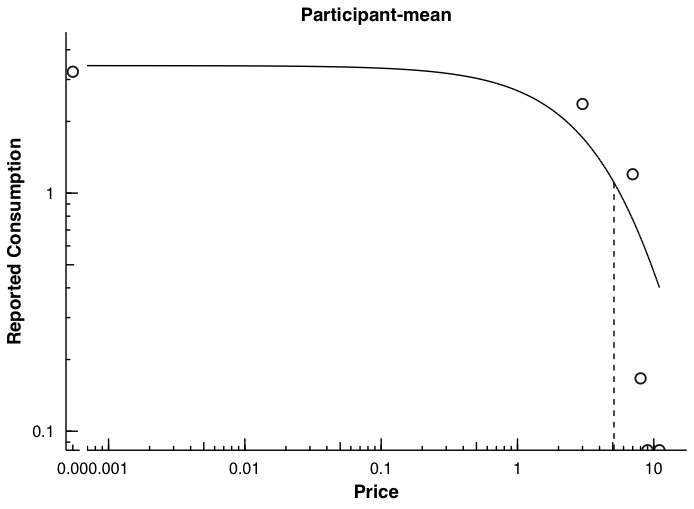 | **Adjusting APT Permutation 8 (*n* = 13)**    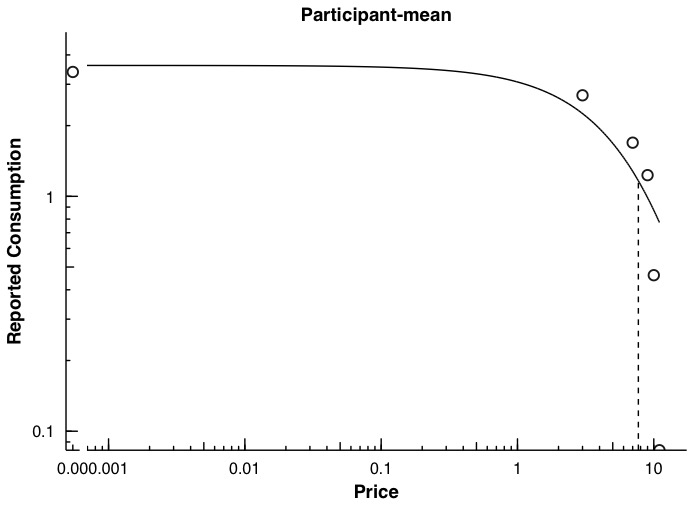 |
| **Adjusting APT Permutation 9 (*n* = 55)**  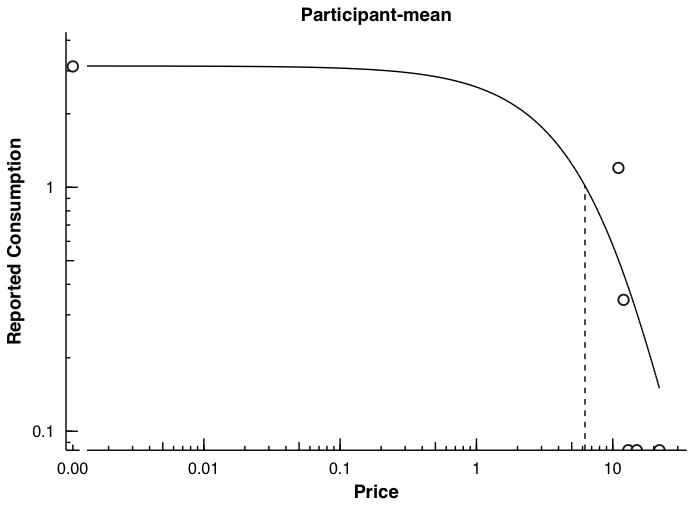 | **Adjusting APT Permutation 10 (*n* = 31)**  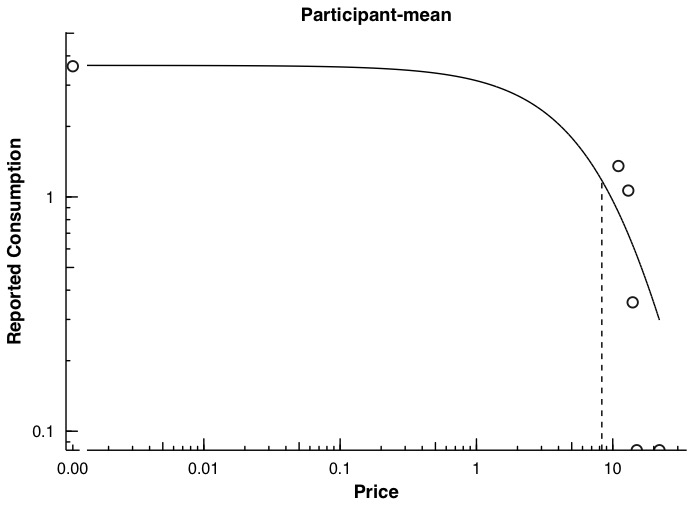 |
| **Adjusting APT Permutation 11 (*n* = 72)**  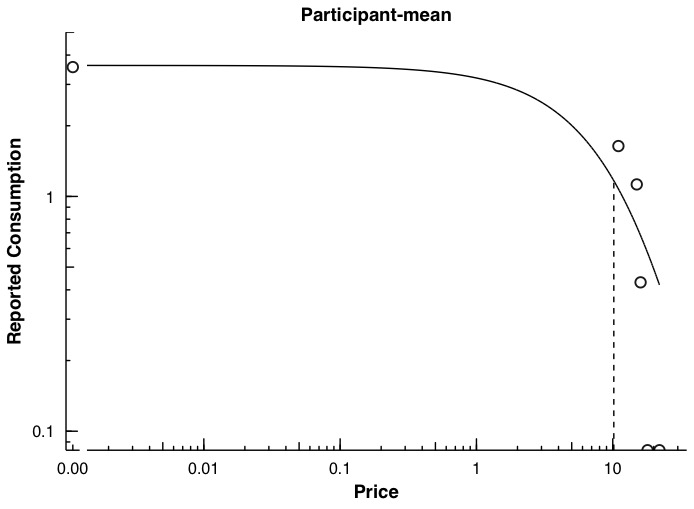 | **Adjusting APT Permutation 12 (*n* = 33)**  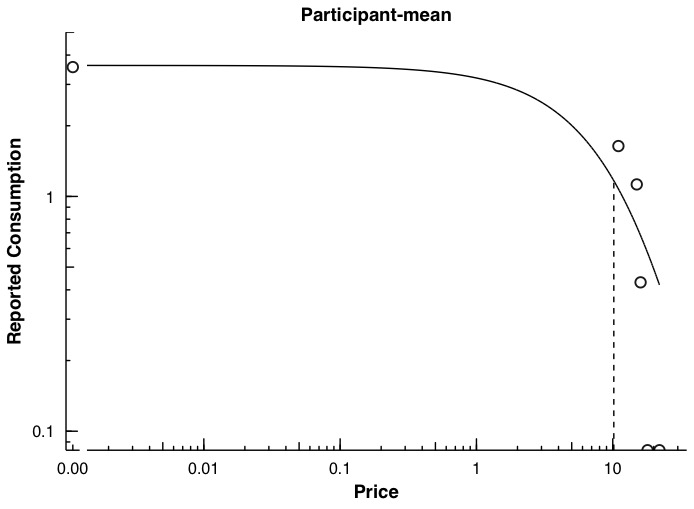 |
| **Adjusting APT Permutation 13 (*n* = 88)**  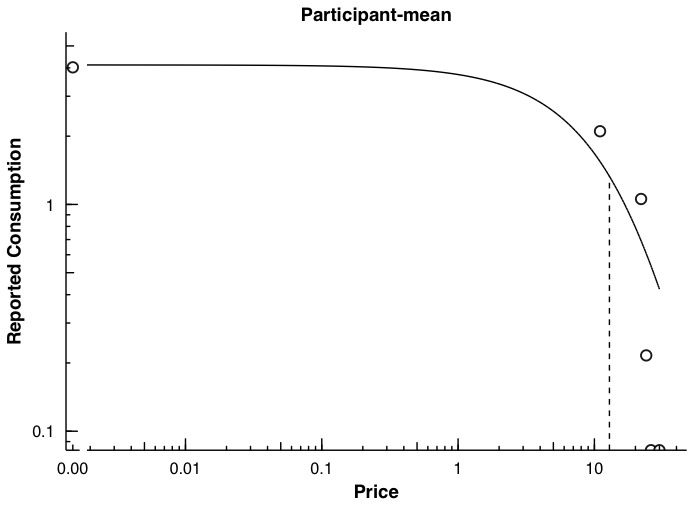 | **Adjusting APT Permutation 14 (*n* = 17)**  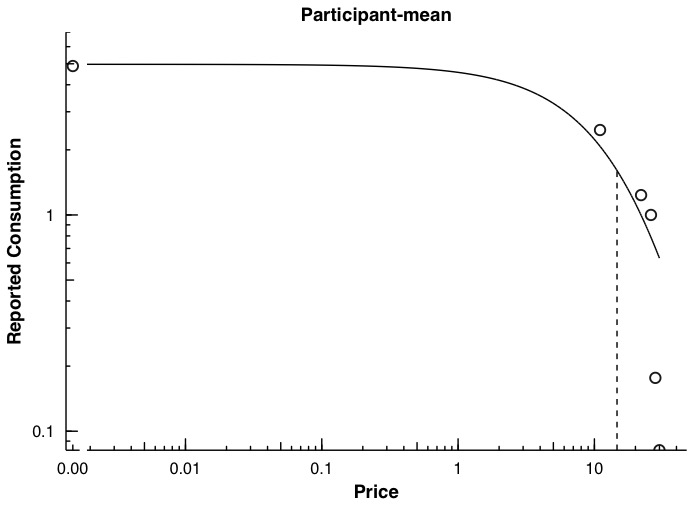 |
| **Adjusting APT Permutation 15 (*n* = 31)**  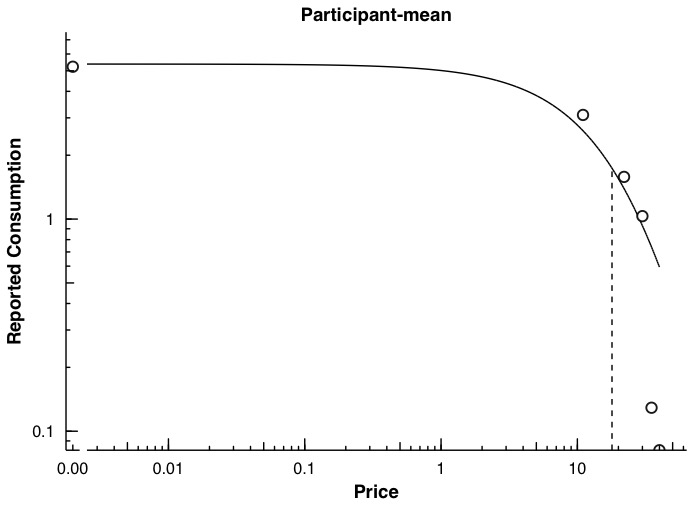 | **Adjusting APT Permutation 16 (*n* = 20)**  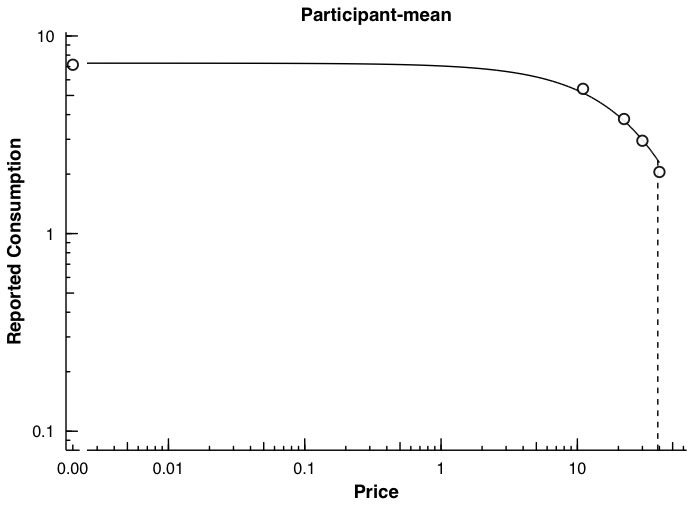 |

*Note.* APT = alcohol purchase task; *n* = number of participants for whom individual demand curves could be estimated (i.e., provided systematic data and reported alcohol consumption at two or more prices) and are thus represented in the plot for a given version of the alcohol purchase task. Both price (*x* axis) and reported consumption (*y* axis) are presented in log scale. A plot for the first permutation of the adjusting alcohol purchase task is omitted, as all participants who were administered this permutation of the task (*n* = 12) reported either zero consumption across all prices or consumption only when alcohol was free, precluding the estimation of individual demand curves. Price sequences corresponding to each adjusting alcohol purchase task permutation are reported in Table S3.
